# Causally Associations of Blood Lipids Levels with COVID-19 Risk: Mendelian Randomization Study

**DOI:** 10.1101/2020.07.07.20147926

**Authors:** Kun Zhang, Yan Guo, Zhuo-Xin Wang, Jing-Miao Ding, Shi Yao, Hao Chen, Dong-Li Zhu, Kun Zhang, Wei Huang, Shan-Shan Dong, Tie-Lin Yang

## Abstract

**Background:** Coronavirus disease 2019 (COVID-19) is a global pandemic caused by the severe acute respiratory syndrome coronavirus 2 (SARS-Cov-2). It has been found that coronary artery disease (CAD) is a comorbid condition for COVID-19. As the risk factors of CAD, whether blood lipids levels are causally related to increasing susceptibility and severity of COVID-19 is still unknown.

**Design:** We performed two-sample Mendelian Randomization (MR) analyses to explore whether dyslipidemia, low density lipoprotein cholesterol (LDL-c), high density lipoprotein cholesterol (HDL-c), triglyceride (TG) and total cholesterol (TC) were causally related to COVID-19 risk and severity. The GWAS summary data of blood lipids involving in 188,578 individuals and dyslipidemia in a total of 53,991 individuals were used as exposures, respectively. Two COVID-19 GWASs including 1,221 infected patients and 1,610 severe patients defined as respiratory failure were employed as outcomes. Based on the MR estimates, we further carried out gene-based and gene-set analysis to explain the potential mechanism for causal effect.

**Results:** The MR results showed that dyslipidemia was casually associated with the susceptibility of COVID-19 and induced 27% higher odds for COVID-19 infection (MR-IVW OR = 1.27, 95% CI: 1.08 to 1.49, *p-value* = 3.18 × 10^−3^). Moreover, the increasing level of blood TC will raise 14 % higher odds for the susceptibility of COVID-19 (MR-IVW OR = 1.14, 95% CI: 1.04 to 1.25, *p-value* = 5.07 × 10^−3^). Gene-based analysis identified that *ABO* gene was associated with TC and the gene-set analysis found that immune processes were involved in the risk effect of TC.

**Conclusions:** We obtained three conclusions: 1) Dyslipidemia is casually associated with the susceptibility of COVID-19; 2) TC is a risk factor for the susceptibility of COVID-19; 3) The different susceptibility of COVID-19 in specific blood group may be partly explained by the TC concentration in diverse ABO blood groups.

## Introduction

Coronavirus disease 2019 (COVID-19) caused by the severe acute respiratory syndrome, coronavirus 2 (SARS-Cov-2) is a global pandemic ^1, 2^. This disease progresses from asymptomatic to acute respiratory distress syndrome and multiple organ dysfunction, and has become a major threat to public health in more than 160 countries ^1, 2^. As of June 26, 2020, there were more than 9.69 million confirmed cases, with total deaths increasing to 487,997 worldwide. Considering the severity of COVID-19, it is urgent to explore the susceptibility factors of COVID-19, which is helpful to develop effective policies and personalized treatments to control the spread of the disease to susceptible groups.

Recent studies ^1, 2^ have found that more than 20% of the confirmed cases had a history of coronary artery disease (CAD). Blood lipids, including low density lipoprotein cholesterol (LDL-c), high density lipoprotein cholesterol (HDL-c), triglyceride (TG) and total cholesterol (TC) are heritable and modifiable risk factors for CAD ^3, 4^. However, whether blood lipids levels are causally related to increasing susceptibility and severity for COVID-19 is still unknown.

Mendelian randomization (MR) is an epidemiological method in which environmental exposure-related genetic variations are used as instrumental variables (IVs) to evaluate the association between exposures and outcomes ^5, 6^. It can avoid the issues of confusion and has been demonstrated as an effective strategy to identify causal effect ^5-10^. In this study, we conducted a two-sample Mendelian randomization study to explore the possible causal associations between blood lipids and COVID-19 susceptibility and severity, and investigate the potential mechanisms underlying the causal effect.

## Methods

A step-by-step workflow in this study is presented in Figure 1.

**Figure 1.**
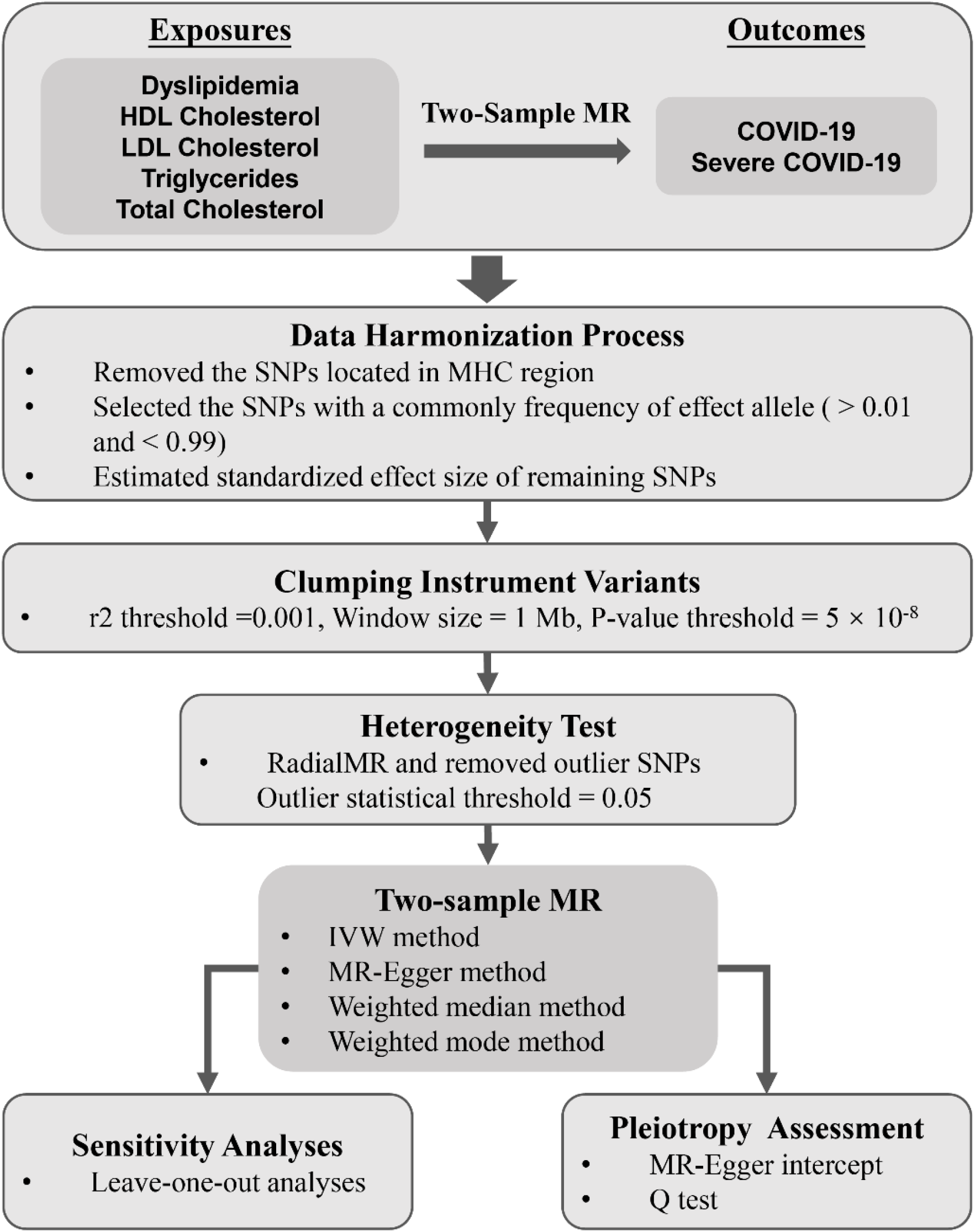
The MR analysis pipeline of the current study.

### Datasets used in this study

#### Blood lipids

We collected data for HDL-c, LDL-c, TG and TC from a published genome-wide association study (GWAS) ^3^ (http://csg.sph.umich.edu//abecasis/public/lipids2013/) involving in a total of 188,578 European-ancestry individuals. GWAS summary data of dyslipidemia was obtained from the Genetic Epidemiology Research on Adult Health and Aging (GERA) (https://cnsgenomics.com/content/data) with 53,991 individuals ^10^.

#### COVID-19

For assessment of associations with COVID-19 risk, we used the latest COVID-19 GWAS results from GRASP database (https://grasp.nhlbi.nih.gov/Covid19GWASResults.aspx). The phenotype used in this GWAS was case/control for COVID-19 infection containing 1,221 positive COVID-19 tests and 4,117 negative tests from UK Biobank individuals (released on June 5, 2020). To explore the causal effect of blood lipids on severity of COVID-19, we also accessed GWAS of severe COVID-19 defined as respiratory failure ^11^ (http://www.c19-genetics.eu/). This GWAS included 1,610 severe COVID-19 patients and 2,205 control participants from Italy and Spain. These two GWASs were all conducted with the correction of age, sex and top 10 principal components.

### Data Harmonization Process

For each exposure GWAS, we performed harmonization process using the following criteria:

1. Remove the SNPs located in major histocompatibility complex (MHC) region.
2. Select the SNPs with a common frequency of the effect allele (EAF) (> 0.01 and < 0.99).
3. Standardize the effect size (β) and standard error (se) for each GWAS data with the function of minor allele frequency and sample size using the following formula ^12^:

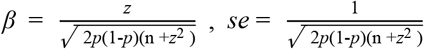

where *z* = β/se from the original summary data, *p* is the minor allele frequency, and n is the total sample size.

### Instrumental variables (IVs) selection

We selected independent and genome-wide significant GWAS SNPs of HDL-c, LDL-c, TG, TC and dyslipidemia by use of the clumping algorithm in PLINK (http://pngu.mgh.harvard.edu/purcell/plink/) ^13^ at a suggestive threshold (r^2^ threshold = 0.001, window size = 1 Mb, *p-value* = 5 × 10^−8^). The 1000 Genomes Project (http://www.internationalgenome.org/) data were used as the reference for linkage disequilibrium (LD) estimation. For each outcome, we then removed outlier pleiotropic SNPs using RadialMR ^14^ with the *p-value* threshold of 0.05. RadialMR ^14^ identified outlying genetic instruments via heterogeneity test (modified Q-statistics). After the removal of pleiotropy, the remaining exposure related SNPs for each outcome as instrumental variables (IVs) were utilized to perform MR analyses.

### MR analyses and pleiotropy assessment

We conducted four complementary two-sample MR methods, including Inverse-Variance Weighted (IVW) method, weighted median method, weighted mode method and MR-Egger method, which make different assumptions about horizontal pleiotropy.

The IVW method assumes balanced pleiotropy ^15^. The pleiotropy is assessed via Cochran’s *Q* statistic and presented as excessive heterogeneity which will inflate the estimate of MR analysis ^16^. The MR-Egger method is based on the assumption which indicate instrument strength independent of the direct effects ^15^. MR-Egger estimates can be also evaluated by the regression dilution I^2^ _(GX)_ ^17^ according to the assumption that no measurement error in the SNP exposure effects. If I^2^ _(GX)_ ^17^ was sufficiently low (I^2^ _(GX)_ < 0.9), the correction analysis was conducted to assess the causal effect by SIMEX. The intercept term of MR-Egger method can used for evaluating the directional pleiotropic effect ^18^. When the intercept is zero or its *p-value* was not significant (*p-value* > 0.05) were considered as non-pleiotropy. Moreover, we also used the Rucker’s Q′ statistic ^19^ to measure the heterogeneity for MR-Egger method. If the difference Q − Q′ is sufficiently extreme with respect to a χ2 distribution with the 1 degree of freedom, we indicated that directional pleiotropy is an important factor and MR-Egger model provides a better fit than the IVW method ^20^. All methods of two-sample MR analyses were measured by TwoSampleMR package in R. For various estimates for different measures, we select the main MR method as following rules:

1. If no directional pleiotropy in MR estimates (Q statistic: *p-value* > 0.05, MR-Egger intercept: intercept = 0 or *p-value* > 0.05, Q – Q’: *p-value* > 0.05), IVW method was used.
2. If directional pleiotropy was detected (MR-Egger intercept: intercept ≠ 0 and *p-value* < 0.05, Q – Q’: *p-value* < 0.05) and *p-value* > 0.05 for the test of Q’, MR-Egger method was used.
3. If directional pleiotropy was detected (MR-Egger intercept: intercept ≠ 0 and *p-value* < 0.05, Q – Q’: *p-value* < 0.05) and *p-value* < 0.05 for the test of Q’, weighted median method was used.

### Sensitivity analysis

Leave-one-out sensitivity analysis was implemented to assess whether any significant results were generated by a specific SNP in IVW models.

### Gene-based and gene-set analysis

MAGMA (https://ctg.cncr.nl/software/magma) ^21^ is commonly used for gene and gene-set analyses based on GWAS and genotype data. In order to explore the association of TC and COVID-19, we implemented MAGMA to identify genes and gene sets in which multiple SNPs show moderate association to TC without reaching the stringent genome-wide significance level.

#### Genome-wide gene-based association study

The genome-wide gene-based association study (GWGAS) is based on the model of multiple linear principal components regression and calculated the gene *p-value* using F-test ^22^. All 19,427 protein-coding genes from the NCBI 37.3 gene definitions were employed for SNPs annotation. We mapped SNPs to genes by a defined window around each gene of 2kb away from the transcription start site (TSS) upstream and 1kb away from the transcription stop site downstream based on human reference assembly (GRCh37 or hg19) ^23^. The GWGAS analysis was performed to quantify the degree of association for each gene to TC and to compute the correlations between genes are estimated according to LD statistics. The LD reference was also from Phase 3 of 1,000 Genomes.

#### Gene-set analysis

The gene-set analysis is built as an independent layer around the gene analysis, while it also on the strength of the gene *p-value* and gene correlation matrix from the previous step in order to compensate for the dependencies between genes ^22^. A total of 7,521 gene sets were derived from Gene Ontology (GO) biological processes and the 186 pathways from Kyoto Encyclopedia of Genes and Genomes (KEGG) as auxiliary files for gene-set analysis, which obtained from the Molecular Signatures Database (MSigDB) version 6.0 (https://www.gsea-msigdb.org/gsea/downloads.jsp).

### Multiple testing correction

We employed false discovery rate (FDR) to address multiple comparisons issue and the adjusted *p-value* < 0.05 was used for judging significance. For MR estimates, we adopted an FDR control procedure for the susceptibility and severity of COVID-19 separately.

## Results

### IVs selection

After data harmonization and clumping, we identified 20 SNPs, 101 SNPs, 79 SNPs, 59 SNPs and 98 SNPs for dyslipidemia, HDL-c, LDL-c, TG and TC, respectively. For each outcome, the final IVs are shown in Table S1.

### Causal effect of dyslipidemia on COVID-19

We evaluated whether dyslipidemia is causally related to COVID-19 firstly. The assessment of pleiotropy is shown in Table S1. Since there was no significant evidence of pleiotropy (all *p value* > 0.05, Table S1), we chose IVW as the main MR method. We found that dyslipidemia was causally associated with the susceptibility of COVID-19 after FDR correction (MR-IVW *p-value* = 3.18 × 10^−3^, FDR = 1.30 × 10^−2^) (Table 1). The estimate of IVW showed that dyslipidemia could raise 27% odds for the infection risk of COVID-19 (MR-IVW OR = 1.27, 95% CI: 1.08 to 1.49) (Figure 2). Besides the IVW method, the weighted median and MR-Egger tests also showed consistent causal associations (MR-Weighted median OR = 1.26, 95% CI: 1.01 to 1.56, *p-value* = 4.00 × 10^−2^; MR-Egger OR = 1.53, 95% CI: 1.12 to 2.10, *p-value* = 2.00 × 10^−2^) (Table 1). However, dyslipidemia had no causally relevance to severe COVID-19 (Table 1 and Figure 2). In sensitivity analyses, the results of leave-one-out permutation didn’t find individual influential SNPs in IVW models (*p-value* < 0.05) (Figure S1).

**Table 1.** Summary of the MR estimates for dyslipidemia, TC, TG, LDL-c and HDL-c to the susceptibility and severity of COVID-19^1^

| Lipid | IVW method |  | Weighted median method |  | Weighted mode-based method |  | MR-Egger method |  | FDR |
| --- | --- | --- | --- | --- | --- | --- | --- | --- | --- |
|  | OR (95%CI) | *p* | OR (95%CI) | *p* | OR (95%CI) | *p* | OR (95%CI) | *p* |  |
| **COVID-19** |  |  |  |  |  |  |  |  |  |
| Dyslipidemia | 1.27(1.08, 1.49) | 0.003 | 1.26 (1.01, 1.56) | 0.040 | 1.22(0.93, 1.61) | 0.170 | 1.53(1.12, 2.10) | 0.020 | **0.013** |
| Total Cholesterol | 1.14 (1.04, 1.25) | 0.005 | 1.23 (1.06, 1.43) | 0.005 | 1.14 (0.99, 1.32) | 0.070 | 1.15 (0.10, 1.33) | 0.061 | **0.013** |
| Triglycerides | 1.18 (0.98, 1.25) | 0.102 | 1.08 (0.90, 1.31) | 0.392 | 1.15 (0.94, 1.41) | 0.179 | 1.15 (0.94, 1.40) | 0.170 | 0.128 |
| LDL Cholesterol | 1.09 (1.00, 1.18) | 0.051 | 1.06 (0.94, 1.20) | 0.358 | 1.06 (0.95, 1.17) | 0.300 | 1.02 (0.91, 1.15) | 0.697 | 0.085 |
| HDL Cholesterol | 1.03 (0.92, 1.15) | 0.646 | 1.09 (0.91, 1.29) | 0.359 | 1.19 (0.98, 1.44) | 0.091 | 1.21 (0.99, 1.49) | 0.073 | 0.646 |
| **Severe COVID-19** |  |  |  |  |  |  |  |  |  |
| Dyslipidemia | 0.83(0.67, 1.02) | 0.071 | 0.81(0.62, 1.07) | 0.139 | 0.83(0.60, 1.14) | 0.267 | 0.69(0.44, 1.06) | 0.112 | 0.132 |
| Total Cholesterol | 0.95 (0.85, 1.06) | 0.366 | 0.98 (0.81, 1.19) | 0.857 | 1.02 (0.84, 1.24) | 0.856 | 1.00 (0.84, 1.20) | 0.993 | 0.366 |
| Triglycerides | 1.13 (0.99, 1.30) | 0.079 | 1.10 (0.91, 1.33) | 0.344 | 1.07 (0.91, 1.27) | 0.420 | 0.99 (0.80, 1.24) | 0.956 | 0.132 |
| LDL Cholesterol | 0.90 (0.81, 1.00) | 0.049 | 0.95 (0.81, 1.12) | 0.551 | 0.94 (0.81, 1.09) | 0.408 | 0.92 (0.80, 1.07) | 0.269 | 0.132 |
| HDL Cholesterol | 0.92 (0.81, 1.04) | 0.185 | 0.88 (0.72, 1.08) | 0.211 | 0.90 (0.72, 1.13) | 0.361 | 0.89 (0.71, 1.12) | 0.334 | 0.231 |

<sup>1</sup> Results described MR estimates derived from the main inverse-variance weighted, MR-Egger, weighted median and weighted mode-based methods for dyslipidemia, HDL-c, LDL-c, TC and TG to the susceptibility of COVID-19 (COVID-19) and the severity of COVID-19 (Severe COVID-19). FDR refers to the *p-value* from IVW method. Values of FDR < 0.05 are marked in italic bold. *p*, *p-value*; CI, confidence interval.

**Figure 2.**
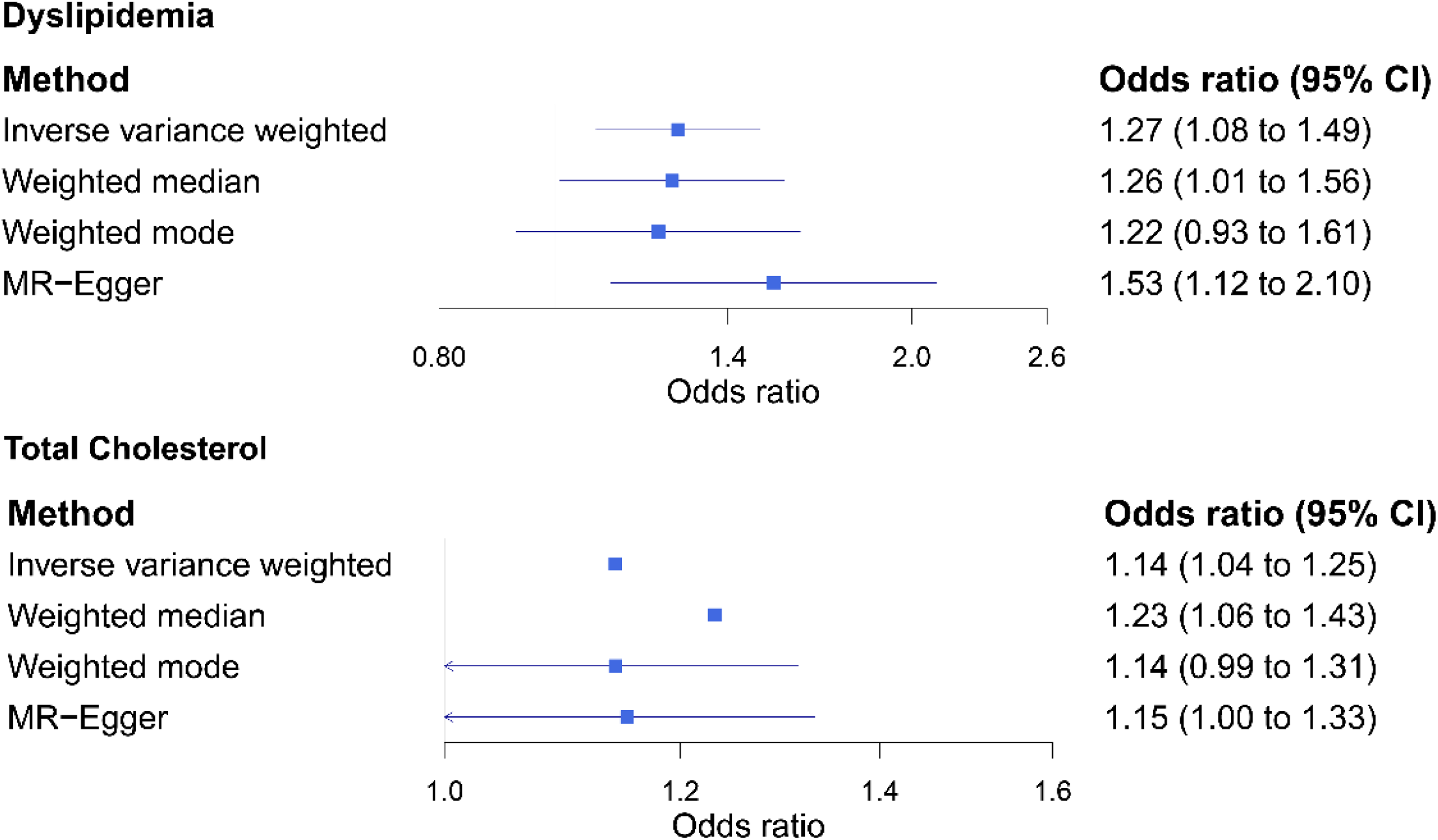
Causal effects of dyslipidemia and TC on COVID-19. Summary of the Mendelian randomization (MR) estimates derived from the main inverse-variance weighted, MR-Egger, weighted median and weighted mode-based methods for dyslipidemia and TC to the susceptibility of COVID-19 (COVID-19).

### Causal effect of blood lipids on COVID-19

We further assessed the causal effects of blood lipids levels, including HDL-c, LDL-c, TC and TG on COVID-19 to identify the specific risk lipid. We didn’t detect any evidence of pleiotropy as well (all *p value* > 0.05, Table S1), thus we still chose IVW as the main MR method. We identified TC was a risk factor for the susceptibility of COVID-19 (MR-IVW *p-value* = 5.07 × 10^−3^, FDR = 1.30 × 10^−2^) (Table 1). As shown in Figure 2, the increasing concentration of TC in blood could induce 14 % higher odds of COVID-19 infection risk (MR-IVW OR = 1.14, 95% CI: 1.04 to 1.25). Besides, the weighted median test also showed a causal association (MR-Weighted median OR = 1.23, 95% CI: 1.06 to 1.43, *p-value* = 5.00× 10^−3^) (Table 1). Leave-one-out analysis indicated that no single SNP was driving the causal estimates (Figure S1). We also measured the relationship between four blood lipids and severe COVID-19. Consistent with dyslipidemia, there was no causal effects for blood lipids-COVID-19 severity pairs.

### Gene-based and gene-set analyses for TC

In our above results, we have discovered the risk effect of TC to the susceptibility of COVID-19. We wonder explain the internal linkage between TC and COVID-19 preliminarily, thus we investigated the potential mechanism of TC by gene-based and gene-set analysis.

#### Gene-based analysis for TC

For gene-based analysis, a total of 17,699 genes which were represented by at least one SNP were identified. After correction for multiple testing, we identified 668 genes linked to TC (Figure 3A and Table S2). It should be noticed that *ABO* was identified to be associated with TC significantly (*p-value =* 6.80 × 10^−11^, FDR *=* 1.43 × 10^−8^). Some observational studies have found that the level of blood lipids was related to ABO blood group and Table 2 lists the detailed information about these observational studies ^24-29^. All of these studies provide a conclusion that TC is higher in A or non-O blood group, but lower in O blood group. On the other side, GWAS on severe COVID-19 has revealed the relationship between ABO blood group locus and COVID-19 ^11^. It has been found a higher risk in blood group A than in other blood group and a protective effect in blood group O, which was coincident with the results of observational investigations based on phenotype ^30-32^. In general, we inferred that the different susceptibility of COVID-19 in specific blood group may be partly explained by the TC levels in diverse ABO blood group (Figure 3B).

**Table 2.** Summary of the observational studies for TC level in different ABO blood group^1^

| Subjects | Cohort | Sample size | ABO blood groups | Mean level of total cholesterol | *p-value* | Description |
| --- | --- | --- | --- | --- | --- | --- |
| Healthy blood donors | Italian | 7,723 | A vs. O | 183.95 ± 0.65 vs.<br>181.31 ± 0.64 (mg/dL) | $1.00 \times 10^{-3}$ | The values of TC and LDL-C are significantly higher in subjects with blood group A compared with those with O blood type <sup>24</sup>. |
| CAD patients | Chinese | 6,476 | Non-O vs. O | 4.93 ± 0.02 vs.<br>4.78 ± 0.03 (mmol/L) | $3.80 \times 10^{-7}$ | Subjects of non-O type had higher levels of TC, LDL-C, and NHDL-c <sup>25</sup>. |
| Adolescents | White | 4,460 | A<sub>1</sub> vs. O | Males: 174.20 ± 0.90 vs.<br>170.70 ± 0.80 (mg/dL)<br>Females: 181.00 ± 1.00 vs.<br>176.10 ± 0.90 (mg/dL) | Male:<br>$5.00 \times 10^{-3}$<br>Female:<br>$3.00 \times 10^{-4}$ | Blood group A with higher serum TC levels in white adolescents <sup>26</sup>. |
| Acute STEMI patients | European | 1,835 | Non-O vs. O | A:180.40 ± 34.50, B:182.10 ± 35.10, AB:180.50 ± 33.80 vs. O:175.30 ± 33.20 (mg/dL) | $2.30 \times 10^{-2}$ | The prevalence of hyperlipidemia, TC, LDL, peak CKMB and no-reflow as well as hospitalization duration were higher in patients with non-O blood groups <sup>27</sup>. |
| FH patients | White | 668 | Non-O vs. O | 9.48 ± 1.69 vs.<br>9.14 ± 1.73 (mmol/L) | $2.00 \times 10^{-2}$ | Total cholesterol was significantly higher in non-O subjects compared to carriers of the O group <sup>28</sup>. |
| Coronary atherosclerosis patients | Chinese | 371 | Non-O vs. O | 5.05 ± 0.07 vs.<br>4.80 ± 0.10 (mmol/L) | $4.10 \times 10^{-2}$ | Subjects of non-O type had higher levels of TC, LDL-C and NHDL-C compared with that of O type <sup>29</sup>. |

<sup>1</sup> Table lists the published studies which demonstrate the TC level is different in diverse ABO blood group. STEMI, ST-elevation myocardial infarction; CAD, coronary artery disease; FH, familial hypercholesterolemia.

**Figure 3.**
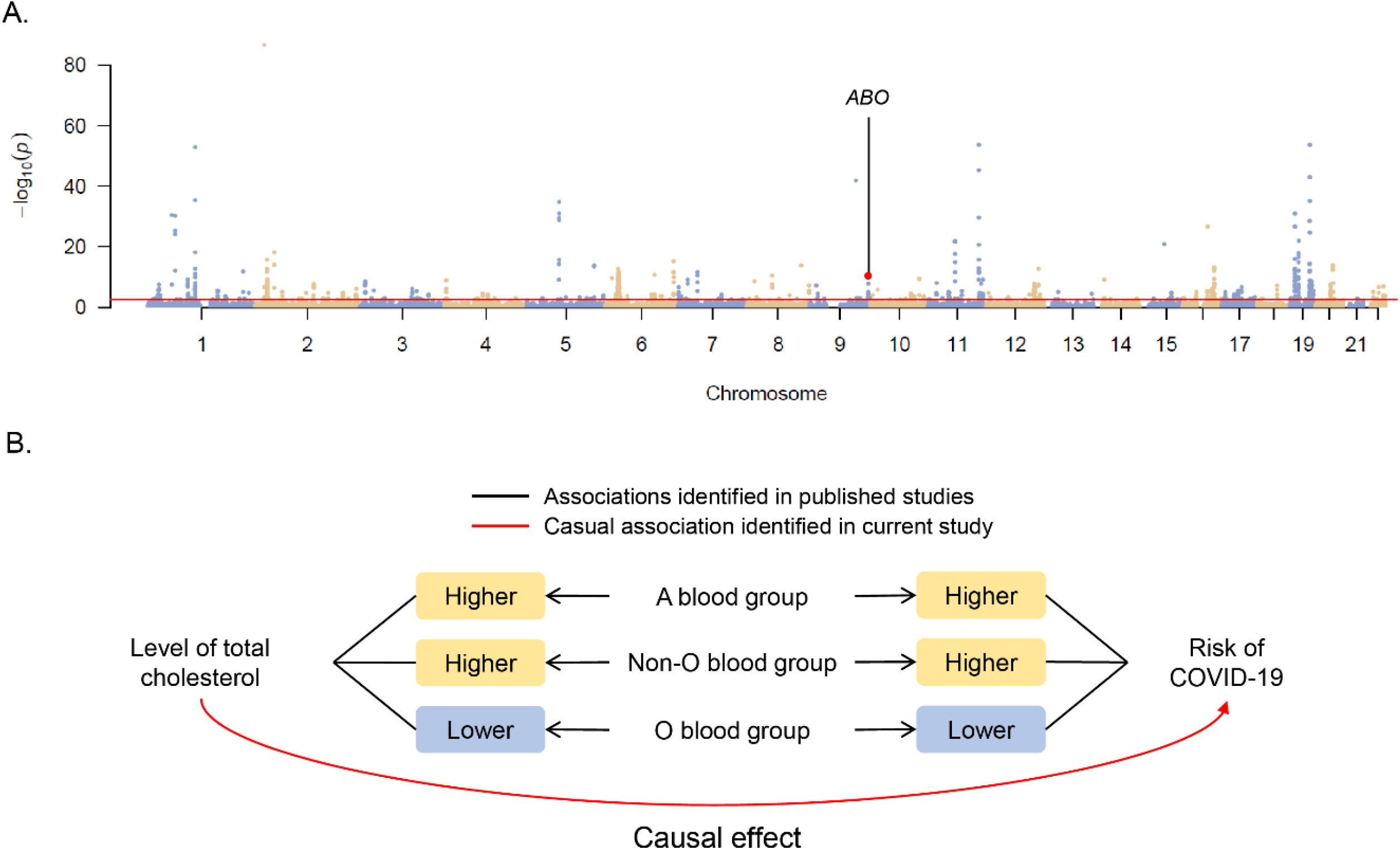
The relationships among the TC level, the susceptibility of COVID-19 and the different ABO blood group. A. Results of gene-based analysis. The dots above the red line represents the genes significantly associated (FDR < 0.05) with TC. The red dot indicates *ABO* gene. B. Schematic diagram displays the relationships among the level of TC, the risk of COVID-19 infection and different ABO blood group. Black lines represent the associations identified in previous studies and red line meansthe causal effect identified in our study.

#### Gene-set analysis for TC

For the GO biological processes, we identified 89 processes significantly associated with TC (FDR < 0.05), which are mostly involved in lipid metabolism (Table S3). For the KEGG pathways, we just identified 2 pathways (Carbohydrate metabolism and Glycan biosynthesis and metabolism) significantly associated with TC after FDR corrections. Besides, we also found 20 KEGG pathways with nominally significant associations with TC (*p-value* < 0.05) (Table S4). In all related processes/pathways, six of them are related to immune response and may participate in the infection and progression of COVID-19, including negative regulation of lymphocyte mediated immunity, interleukin-10 biosynthetic process, interferon-β (INF-β) biosynthetic process, Fc gamma R-mediated phagocytosis, antigen processing and presentation and primary immunodeficiency. The dysfunction of immunity would induce poor immune response for SARS-CoV-2 infection, which cause lung and systemic pathology ^33, 34^.

## Discussion

In this study, we implemented two-sample MR analyses to explore the possible causal associations between blood lipids and COVID-19. We have found potential causal effects of dyslipidemia and blood TC on the infected risk of COVID-19.

To explain the potential influence of TC on COVID-19, we explored the TC-related genes and gene sets. It is notable that *ABO* gene performs quite strong relevance to TC, which was also reported by previous GWAS of TC ^3^. Besides, some observational studies have found that the blood lipids level was related to ABO blood group. The higher level of TC was found in non-O blood group and was significantly associated with an increased prevalence of CVD ^24-29^. In addition, The GWAS of severe COVID-19 has identified the association signal at ABO blood group locus ^11^. Based on the blood-group-specific analysis, they observed a higher risk of COVID-19 in blood group A than in other blood group and a protective effect in blood group O, which was coincident with the results of observational investigations based on phenotype ^30-32^. In summary of these results, we inferred that the different susceptibility of COVID-19 in specific blood group may be partly explained by the TC concentration in diverse ABO blood groups.

The result of gene-set analysis identified a total of 89 biological processes and 20 pathways associated with TC. Besides the processes/pathways related to lipid metabolism, six processes/pathways belonged to immune system were shown moderate associations with TC, including three pathways related to immunity and immunodeficiency (negative regulation of lymphocyte mediated immunity, antigen processing and presentation, primary immunodeficiency) and other three processes involved in immune cytokines and phagocytosis (interleukin-10 biosynthetic process, interferon-β biosynthetic process, Fc gamma R-mediated phagocytosis). The biosynthetic process of interleukin-10 (IL-10) and interferon-β (INF-β) can produce the cytokines with pleiotropic effects in immunoregulation and inflammation, which will destroy host immune response if the biosynthesis is broken. The phagocytosis is an essential role in host-defense mechanisms and Fc gamma receptors can recognize foreign extracellular materials and initiate a variety of signals to start immune process. As a disease caused by the SARS-Cov-2, COVID-19 is in close and direct touch with immunity ^33^. Therefore, we raise a hypothesis that the risk effect of TC on COVID-19 may be mediated by the dysfunction of immune system.

This is the first study to characterize the potential causality of blood lipids for the susceptibility and severity of COVID-19 using two-sample MR design rather than observational and perspective studies based on conventional association analysis. The limitations of the current study should be addressed. Due to the limitation of data resource, our findings are based on European cohort which cannot represent the universal conclusions for other ethnic groups. In addition, the potential mechanism of the risk effect for TC was discussed superficially, which needed to carry out further investigation to get more data support and further experimental verification.

In summary, we carried out a two-sample MR design for blood lipids and COVID-19, and obtained following conclusions: 1) Dyslipidemia is causally associated with the susceptibility of COVID-19; 2) The higher total cholesterol level will increase the susceptibility of COVID-19; 3) The different susceptibility of COVID-19 in specific blood group may be partly explained by the TC concentration in diverse ABO blood groups; 4) The risk effect of total cholesterol on COVID-19 may be mediated by the dysfunction of immunity.

## Data Availability

All data generated or analyzed during this study are included in this published article or in the data repositories listed in References.

## Acknowledgements

This study is supported by National Natural Science Foundation of China (31871264, 31970569); Zhejiang Provincial Natural Science Foundation of China (LGF18C060002); Shannxi Provincial key research and development plan (2019ZDLSF01-09) and the Fundamental Research Funds for the Central Universities (xzy032020039, xzy032020023).

## Supporting information

**Figure S1**. Scatter plot and leave-one-out analysis plot for dyslipidemia and total cholesterol (COVID-19).

**Table S1**. Assessment of pleiotropy for dyslipidemia and blood lipids to COVID-19 and severe COVID-19.

**Table S2**. Summary of 243 genes significantly associated (FDR < 0.05) with total cholesterol.

**Table S3**. Summary of 89 biological processes significantly associated (FDR < 0.05) with total cholesterol.

**Table S4**. Summary of 20 pathways associated (*p-value* < 0.05) with total cholesterol.

